# Barriers to surgical care delivery are harming our planet: a case for decentralized provider services

**DOI:** 10.64898/2026.06.30.26354345

**Authors:** Gabriella Y. Hyman, Ramya Reddy, Taylor Wurdeman, Ralph P. Crew, Mark G. Shrime

**Author notes:** **Corresponding author:** Gabriella Y. Hyman, MBBCh, Dip Obst, MPH.

## Abstract

**Background:** Surgical care centralization in the U.S. delays access and increases carbon emissions. Global targets suggest patients live within 2-hours of a surgical facility. This study quantifies the environmental impact of travel for cataract surgery in rural Michigan and models the potential emissions reductions from decentralizing surgical and follow-up services.

**Methods:** A retrospective, cross-sectional study analyzed electronic medical records from a rural Michigan ophthalmology practice (March–November 2023). We calculated travel distances using population-weighted centroids and estimated emissions using U.S. Department of Energy vehicle data. A k-means clustering model optimized additional facility placement, and a gradient analysis identified optimal numbers for decentralization points, for emissions reductions.

**Results:** The 920 patients traveled a median of 55.45 km (IQR: 43.33–88.20 km) for surgery and 55.07 km (IQR: 43.54–87.82 km) for follow-up visits, generating Total Surgical Access Emissions (TSAE) of 57,168 kgCO (median of 59.20 kgCO ; IQR: 32.31–81.87) under the centralized model. The k-means decentralization model and gradient analysis identified 7 hospitals and 9 clinics, respectively, as the optimal expansion points, reducing emissions by 34.07% (19,475 kgCO saved) and 39.52% (22,590 kgCO saved). The Surgical Access Carbon Impact (SACI) model demonstrated that achieving two-hour access to clinic services reduced excess emissions by 54.7%. Sensitivity analyses using fuel-efficient vehicles (Toyota Prius and Tesla Model 3) or reducing follow-up visit frequency reduced emissions by 54.03% (30,888 kgCO) and 25.83% (14,768 kgCO), respectively.

**Conclusion:** Decentralizing surgical services in rural U.S. settings could cut travel-related emissions by up to 40%, significantly reducing healthcare-related carbon footprints while improving timely access to care. The SACI metric provides a novel framework for integrating environmental sustainability into U.S. health policy and service planning.

## Introduction

Timely access to surgical care is essential for averting avoidable disability and death for both emergency and elective surgical conditions [1]. However, access remains limited globally, including in rural regions of high-income countries such as the United States. The concept of “surgical deserts,” introduced by the American College of Surgeons in 2009, highlights areas with inadequate surgical coverage that require patients to travel long distances to receive care [2]. These travel demands create significant barriers in terms of timely access and impose financial costs on patients and health systems [3]. In 2015, the Lancet Commission on Global Surgery (LCoGS) set a benchmark stating that the entire population should live within two hours of a facility capable of providing surgical care [1]. This access indicator has since informed national surgical plans globally [4], yet it does not account for the environmental impact of long travel distances.

Centralized care models-including centers of excellence-consolidate resources and expertise, but frequently overlook the access challenges imposed on rural populations [5,6,7]. These models impose travel burdens, leading to prolonged journey times, delays in care, and increased out-of-pocket expenses, all of which exacerbate health inequities and contribute to worse outcomes for rural patients [8,3]. Travel for care also carries environmental implications: long-distance travel is a key driver of greenhouse gas (GHG) emissions [9]. While alternatives such as virtual follow-up are suitable for some conditions, surgical procedures and their postoperative care generally require in-person attendance, compounding the environmental cost of centralized delivery.

The environmental stakes are considerable. The U.S. healthcare sector accounts for approximately 10% of national GHG emissions [10]. Patient healthcare-related travel alone generates an estimated 35.7 megatons of CO -equivalent annually in the United States, representing 6.4% of total healthcare-sector emissions [12]. Rural patients, who face longer distances, generate higher per-trip emissions than urban counterparts [12]. Yet the carbon cost of surgical care delivery architecture-the geographic distribution and organizational structure of services-has received little empirical attention.

This study quantifies the effect of decentralizing initial and follow-up surgical care on GHG emissions in rural America, using cataract surgery as a representative model. We hypothesize that decentralization of surgical services reduces patient travel-related carbon emissions.

## Methods and Materials

### Overview

This retrospective, cross-sectional study was conducted at a single ophthalmology practice located in a rural area of Michigan, USA. The practice serves a catchment population of approximately 40,000 residents, with no other ophthalmic practices present in the surrounding four counties. The study includes all patients who traveled to the practice and underwent elective cataract surgery (intraocular lens surgery - IOL) at a hospital located in the same hospital complex, from March to November 2023. An IRB waiver was obtained as this was not human subjects (IRB Protocol: 24-1130). We followed STROBE reporting guidelines for observational studies.

### Data Handling and Analysis

Data were collected from the electronic medical records (EMR) of the practice for the following variables: procedure type, date of procedure, number of postoperative visits, and patient residence zip codes. In this study, all procedures took place at the same facility and all postoperative visits occurred at the same clinic. Calculated variables included distance traveled (km) and drive time (minutes). Distance traveled was estimated as the round-trip from the population-weighted centroid of the zipcode where the patient resides to the clinic or hospital, based on geographic data from Open Street Maps (OSM). The OSRM function in R used Dijkstra’s algorithm, optimized with contraction hierarchies, as a way-finding algorithm considering road classification and speed (motorways: 100 km/h, trunk roads: 90 km/h, residential: 50 km/h for defined paths). ArcGIS Online was used for geographic visualization. All calculations were performed using R Studio (version 2024.04.2+764).

### Carbon Emission Calculations

Carbon emissions (in kgCO) were estimated using data on vehicle types, fuel efficiency, and emission factors. We obtained vehicle data in Michigan from publicly available sources to assume of the most commonly registered car in the state - the Ford F-150 Series SUV (Michigan RMV). In addition, we looked at the impact of driving the most fuel-efficient vehicles in Michigan, namely the Toyota Prius and Tesla Model 3 respectively. All fuel economy data and corresponding greenhouse gas emission (GHG) factors (in kilograms of CO per mile) were obtained for model year 2022 from the US Department of Energy.

The total carbon emissions per patient, Total Surgical Access Emissions (TSAE) in kgCO□, were calculated using the following formula:

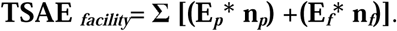

Where: E□(kgCO□emitted) = (distance traveled in km for visit i) × GHG factor (kgCO□/km); E□= emissions for the procedure visit; E□= emissions for a follow-up visit; n□= number of procedures; n□= number of follow-up visits.

### Decentralized Care Model

A modified k-means clustering approach was used to optimize the placement of hypothetical additional hospital and/or clinic locations. This method added *n* additional locations to the catchment area. The initial *n* additional starting points were randomly selected amongst the 47 population-weighted zipcode centroids, with small random offsets to their coordinates. Each population-weighted centroid was assigned to the closest location and the total distance calculated. Each *n* point’s location was recalculated according to a linear weighted average of the longitude and latitude positions. New coordinates’ were weighted by patient count per zipcode in the assigned cluster. The total distance was recalculated iteratively until convergence, which was defined as a < 1% change in total distance traveled. Recalculated points outside the polygon of contiguous zipcodes were snapped to the nearest point within the polygon. The end-point was set where an asymptote was observed in the decentralization model’s trajectory, suggesting that further decentralization yielded minimal additional impact. For each decentralized model, a Wilcoxon-rank-sum was used to compare the reduction of carbon emissions to the centralized model.

The inflection point for optimal reduction in total carbon emissions, based on *n* additional locations, was defined as the value of *n* where the first derivative was positive or zero, or by a change in the sign of the second derivative if the first condition was not met.

To assess the combined effect of decentralizing both surgical and postoperative care, we analyzed total carbon emissions across permutations of n additional hospital and clinic locations. A gradient analysis was performed to calculate the rate of change in total carbon emissions using the Carbon Emissions Saving Rate (CESR):

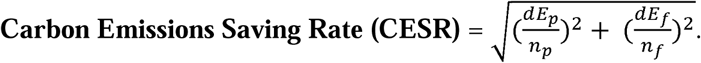

### Surgical Access Carbon Impact (SACI)

We proposed the Surgical Access Carbon Impact (SACI) metric to link the impact of achieving timely access according to global safety targets, with changes in carbon emission. We set sequential timeliness thresholds at 15, 30, 40, 60, 90 and 120 minutes and calculated the excess carbon emissions for patients living outside the acceptable drivetime perimeter. We tested for significant differences in excess emissions at the different drivetime thresholds using a Kruskall-Wallis test.

## Results

### Participants

A total of 920 patients received a single cataract surgical procedure. Four patients were excluded for miscoded zip codes. The 916 (99.6%) included patients, from 47 distinct zip codes, attended a total of 1352 (median per patient = 1; IQR per patient = 1 - 2) follow-up visits, with 6 patients lost to follow-up (Table 1). The hospital and clinic were located in the zip code with the most follow-up visits (n= 240) (Figure 1). The complete list of zipcodes with number of patient residents and postoperative visits is found in Appendix 1.

**Figure 1.**
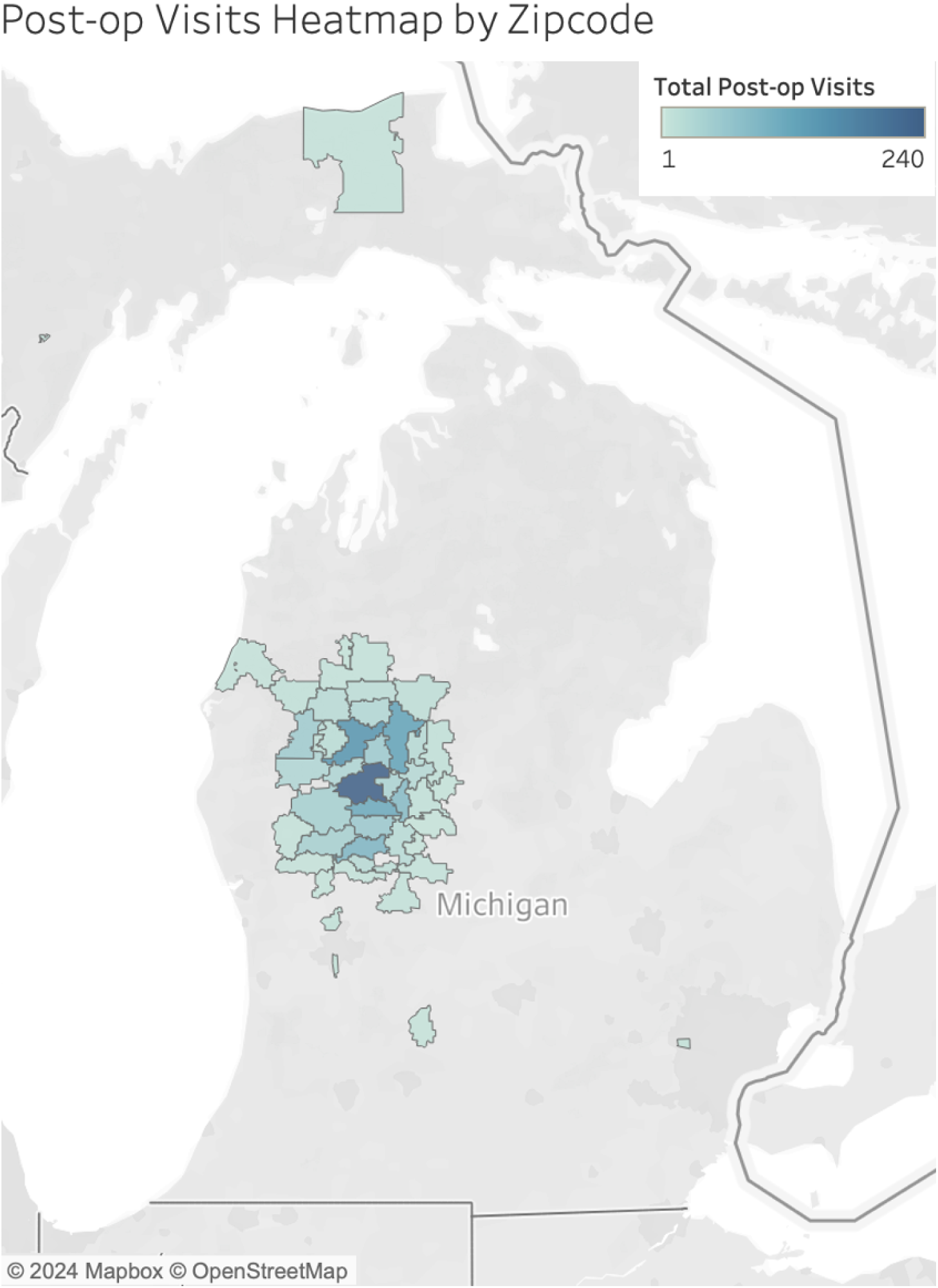
Postoperative Visits per Zip Code.

**Table 1.** Distribution of Post-Operative Visits by Patient and Zipcode.

| Postoperative Visits<br>per Patient | Percentage of<br>Patients | Unique Patient<br>Zipcodes | Median Distance Traveled per<br>Postoperative Visit Roundtrip (km) |
| --- | --- | --- | --- |
| 0 | 0.66% (n = 6) | 5 | 0 |
| 1 | 58.30% (n = 534) | 42 | 55.07 |
| 2 | 34.93% (n = 320) | 42 | 55.06 |
| 3 | 5.35% (n = 49) | 19 | 79.47 |
| 4 | 0.55% (n = 5) | 3 | 0.55 |
| 5 | 0.11% (n = 1) | 1 | 55.06 |
| 6 | 0.11% (n = 1) | 1 | 97.18 |

### Travel Distances and Drivetime

Patients traveled a median of 55.45km (IQR = 43.33 - 88.20 km; range = 1.05-1134.67 km), equivalent to a median travel time of 57.64 minutes (IQR = 42.12 - 87.14 minutes), per surgery. The median travel distance for a single postoperative visit was 55.07 km (IQR = 43.54 - 87.82 km). This corresponded to a median drive time of 56.37 minutes (IQR = 42.02 - 86.20 minutes).

### Carbon Impact of the Centralized Model

Under the centralized care model, the median emissions per procedure and postoperative visit were 20.63 kgCO (IQR = 16.11 - 32.81 kgCO) and 20.48 kgCO (IQR = 16.20 - 32.67 kgCO), respectively. The cumulative emissions for all postoperative visits were 34089.03 kgCO (median = 32.39; IQR = 16.94 - 41.02 kgCO). Total emissions from both the procedure and postoperative visits were 57,167.62 kgCO (median = 59.20; IQR = 32.31 - 81.87 kgCO) (Table 2; Figure 2).

**Figure 2.**
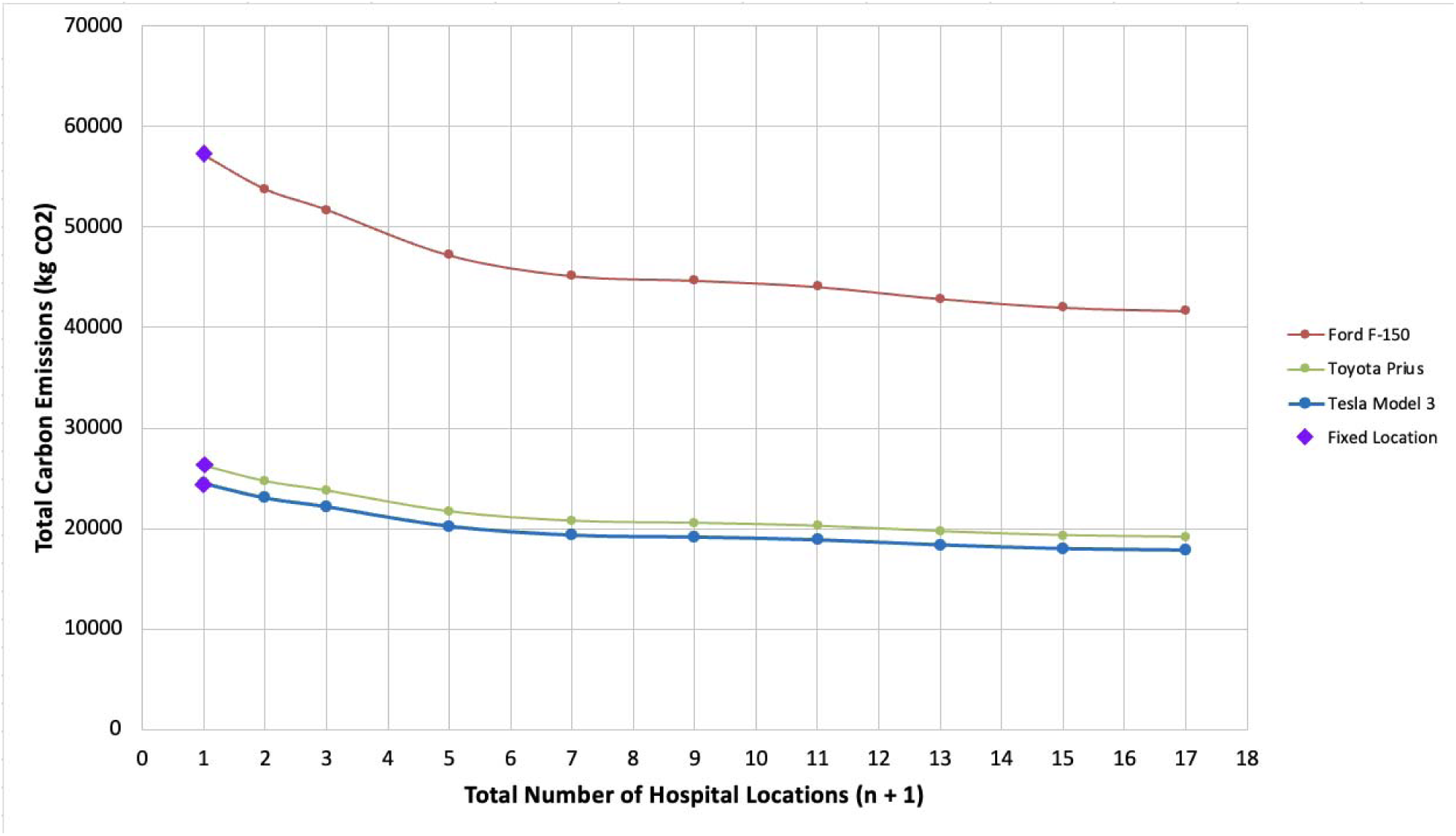
The Impact of Decentralization of Surgical Care on Total Carbon Emissions.

**Table 2.** Total Carbon Emission based on Centralized and Decentralized Surgical Care.

| Number of Hospital Locations | Total Carbon Emissions* kgCO<sub>2</sub> (Median; IQR) | Toyota Prius kgCO<sub>2</sub> (Median; IQR) | Tesla Model 3 kgCO<sub>2</sub> (Median; IQR) |
| --- | --- | --- | --- |
| Fixed Hospital (1)** | 57,167.62 (59.20; 32.31–81.87) | 26,278.66 (27.21; 14.86–37.63) | 24,499.08 (25.38; 13.84–35.09) |
| 2 | 53,777.39 (48.51; 32.31–79.54) | 24,720.25 (22.30; 14.86–36.58) | 23,044.42 (20.78; 13.84–34.07) |
| 3 | 51,734.32 (48.51; 32.31–69.06) | 23,781.10 (22.30; 14.86–31.74) | 22,170.71 (20.78; 13.84–29.59) |
| 5 | 47,208.30 (41.11; 32.31–61.75) | 21,700.59 (18.90; 14.86–28.38) | 20,226.00 (17.61; 13.84–26.46) |
| 7 † | 45,143.06 (39.75; 23.48–61.37) | 20,751.25 (18.27; 10.79–28.17) | 19,336.44 (17.04; 10.07–26.29) |
| 9 | 44,690.36 (40.89; 28.86–61.37) | 20,543.15 (18.80; 13.27–28.17) | 19,158.91 (17.53; 12.37–26.29) |

| Number of Hospital Locations | Total Carbon Emissions*<br>kgCO<sub>2</sub> (Median; IQR) | Toyota Prius kgCO<sub>2</sub><br>(Median; IQR) | Tesla Model 3 kgCO<sub>2</sub><br>(Median; IQR) |
| --- | --- | --- | --- |
| 11 | 44,071.04 (40.21; 27.51–61.37) | 20,258.46 (18.48; 12.65–28.17) | 18,896.00 (17.22; 11.79–26.29) |
| 13 | 42,881.70 (40.89; 25.18–61.37) | 19,711.75 (18.80; 11.57–28.17) | 18,377.31 (17.53; 10.79–26.29) |
| 15 | 42,015.40 (38.49; 23.40–49.74) | 19,313.53 (17.69; 10.76–22.88) | 18,019.91 (16.50; 10.02–21.32) |
| 17 | 41,681.71 (39.13; 22.93–59.66) | 19,160.14 (17.99; 10.55–27.42) | 17,869.76 (16.76; 9.82–25.58) |

Note. †Inflection point. \*Total carbon emissions include emissions from the initial surgical procedure and all postoperative visits. \*\*Centralized care model: single fixed clinic and hospital location..

### Decentralization of Surgical Care

With one additional random hospital location, total emissions decreased by 5.93% to 53,777.39 kgCO (Table 2) (p = 0.01). At the inflection point of 6 additional hospital locations (n = 7), total emissions decreased by 21.03%, saving 12,024.56 kgCO (p < 0.05). With each additional hospital, total emissions reduced by 967.87 kgCO2 (1.05 kgCO2 per patient) (Figure 3). As the number of hospital locations increased, more peripheral zip codes experienced carbon savings, with a median decrease of 17.44% in carbon emissions per zip code at the inflection point (n = 7).

**Figure 3.**
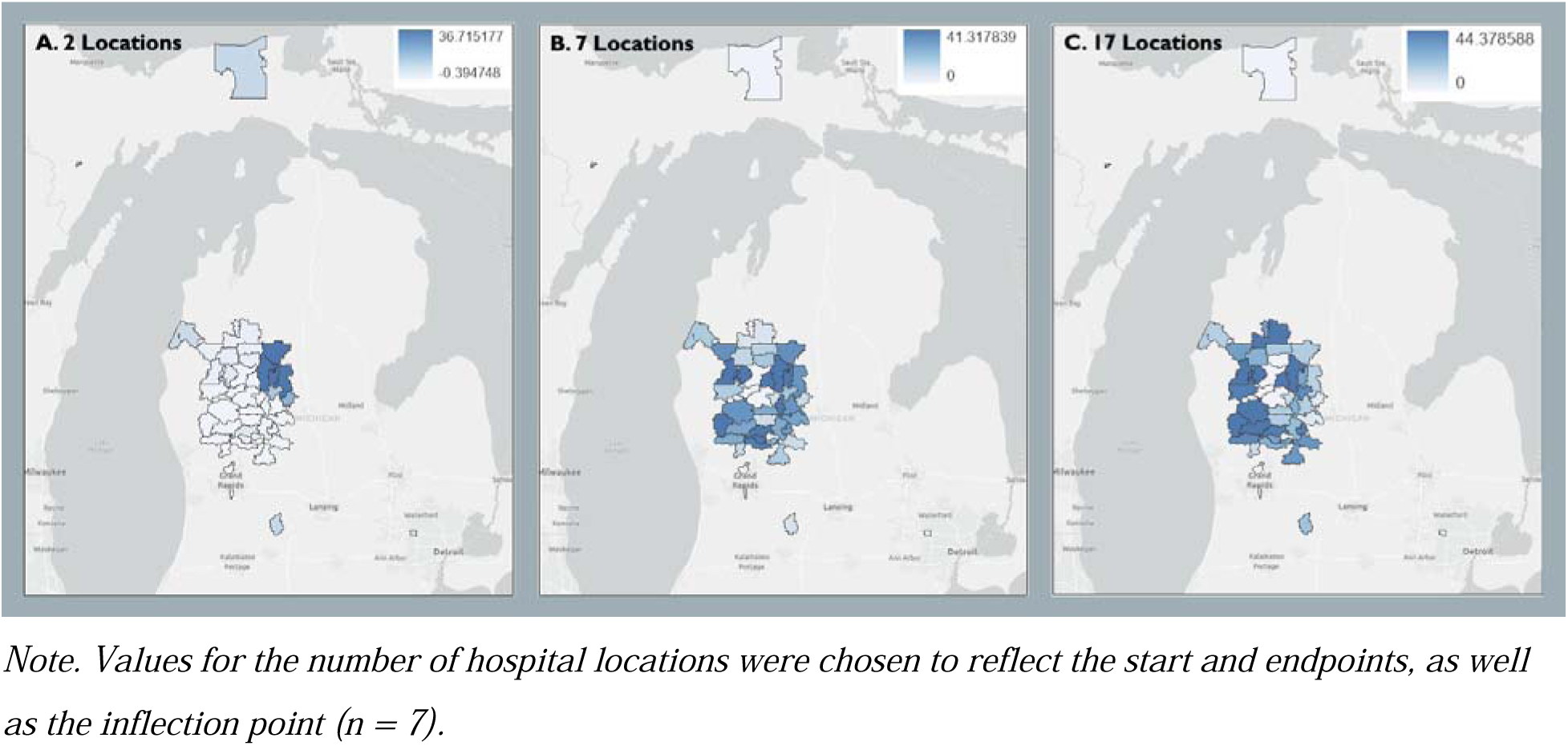
Decentralized Surgical Care: Heatmap of Percent Change in Total Carbon Emissions.

### Decentralization of Postoperative Care

One additional clinic location decreased total emissions by 6775.46 kgCO (11.8%) (p < 0.05), with each additional clinic decreasing total emissions by 1411.91 kgCO (1.54 kgCO per patient) (Table 3; Figure 4). At the inflection point of 8 additional clinic locations (n = 9), total emissions decreased by 34.07%, saving 19475.02 kgCO (p < 0.05) (Figure 5). As the number of clinic locations increased, more peripheral zip codes experienced carbon savings, with a median decrease of 30.40% in carbon emissions per zip code at the inflection point (n = 9).

**Figure 4.**
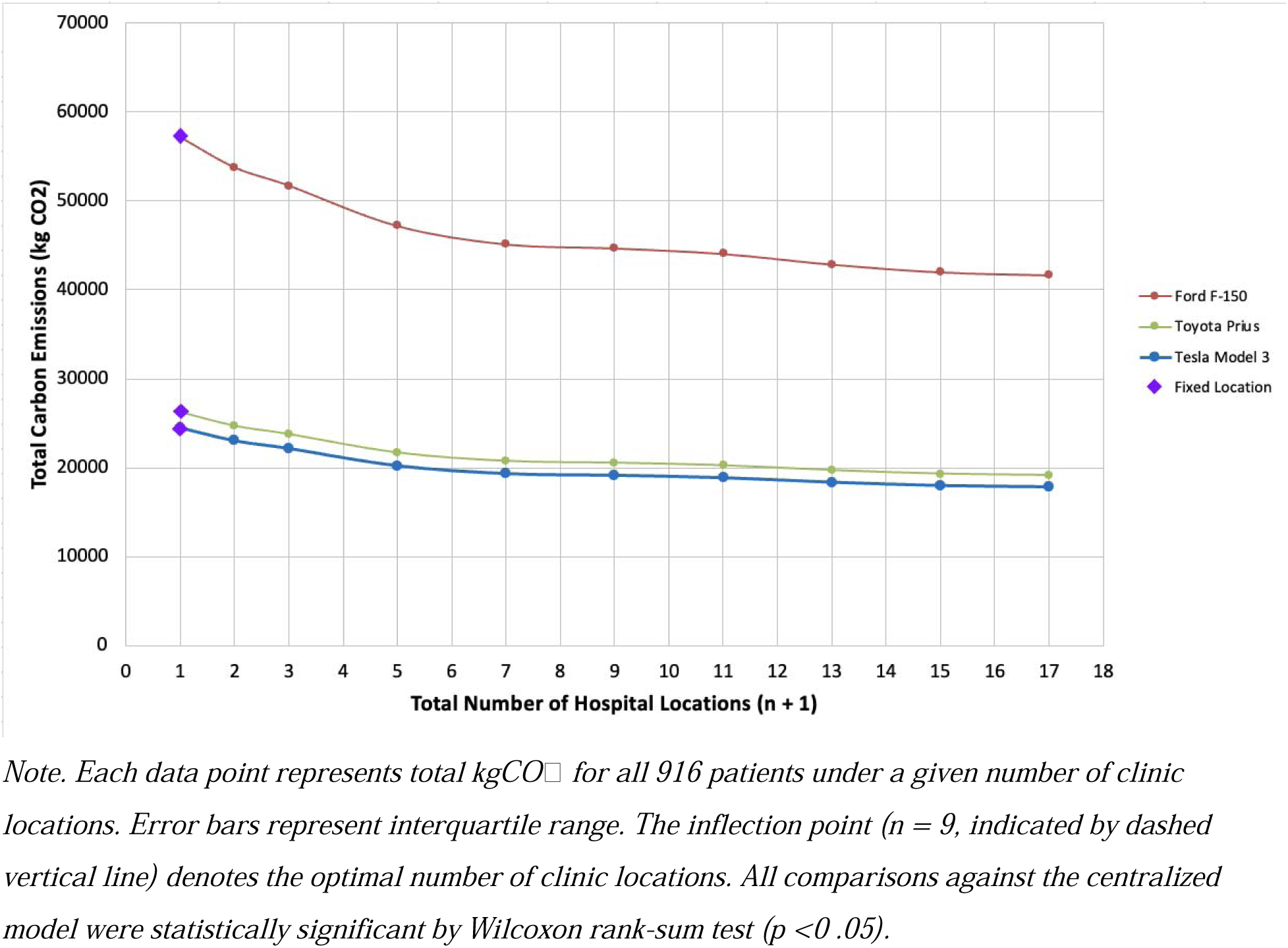
The Impact of Postoperative Decentralization on Total Carbon Emissions.

**Figure 5.**
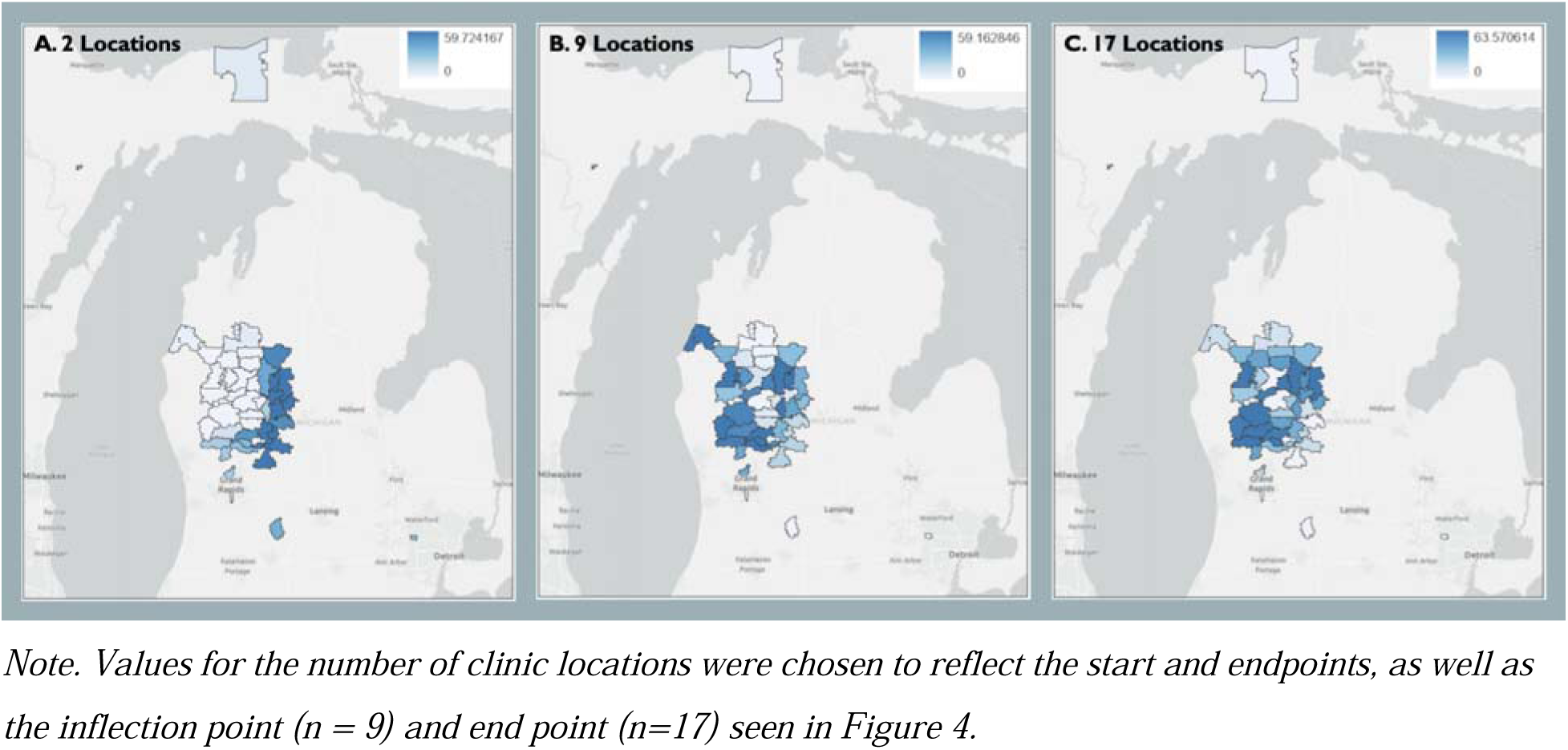
Decentralized Postoperative Care: Heatmap of Percent Change in Total Carbon Emissions.

**Table 3.** Total Carbon Emissions Based on Centralized and Decentralized Postoperative Care.

| Number of Clinic Locations | Total Carbon Emissions* kgCO<sub>2</sub> (Median; IQR) | Toyota Prius kgCO<sub>2</sub> (Median; IQR) | Tesla Model 3 kgCO<sub>2</sub> (Median; IQR) |
| --- | --- | --- | --- |
| Centralized Clinic (1)** | 57,167.62 (59.20; 32.31–81.87) | 26,278.66 (27.21; 14.86–37.67) | 24,511.39 (25.38; 13.84–35.09) |
| 2 | 50,392.16 (48.51; 32.31–65.62) | 23,164.14 (22.30; 14.86–30.17) | 21,606.32 (20.80; 13.84–28.13) |
| 3 | 46,179.77 (40.89; 32.31–61.47) | 21,227.80 (18.80; 14.86–28.26) | 19,800.20 (17.53; 13.84–26.34) |
| 5 | 42,502.71 (40.89; 32.20–58.14) | 19,537.54 (18.80; 14.79–26.72) | 18,223.61 (17.53; 13.80–24.91) |
| 7 | 39,317.55 (37.45; 23.32–49.24) | 18,073.39 (17.21; 10.72–22.64) | 16,857.93 (16.06; 10.00–21.10) |
| 9 † | 37,692.60 (37.47; 22.44–46.37) | 17,326.44 (17.22; 10.31–21.30) | 16,161.21 (16.07; 9.62–19.99) |
| 11 | 36,896.15 (36.21; 16.94–47.58) | 16,960.33 (16.64; 7.79–21.89) | 15,819.72 (15.53; 7.26–20.57) |
| 13 | 36,365.68 (38.61; 24.15–45.21) | 16,716.48 (17.75; 11.10–20.78) | 15,592.27 (16.55; 10.35–19.53) |
| 15 | 34,642.93 (33.63; 20.31–41.78) | 15,924.57 (15.46; 9.34–19.20) | 14,853.62 (14.42; 8.71–18.04) |
| 17 | 34,577.03 (34.75; 20.36–41.72) | 15,894.28 (15.97; 9.36–19.17) | 14,825.37 (14.90; 8.72–18.02) |

*Note. CE = carbon emissions.\*Total carbon emissions include emissions from the initial surgical procedure and all postoperative visits. Comparisons at each step were made against the centralized model using a Wilcoxon rank-sum test; all comparisons were statistically significant ( $p < .05$ ).*
*\*\*Centralized care model: single fixed clinic and hospital location. †Inflection point. See Table A2 for sensitivity analysis accounting for patients lost to follow-up.*

### Combined Decentralized Model

Under the combined decentralized model at the inflection points of 7 hospitals and 9 clinics, total carbon emissions decreased by 55.1% to 25,668.04 kgCO (median: 18.23; IQR: 9.24-34.00 kgCO) (Figure A1). Compared with individual decentralization strategies, the combined model further reduced emissions relative to hospital-only decentralization by 43.14% (p<0.05) and relative to clinic-only decentralization by 31.90% (p<0.05) (Figure 6).

**Figure 6.**
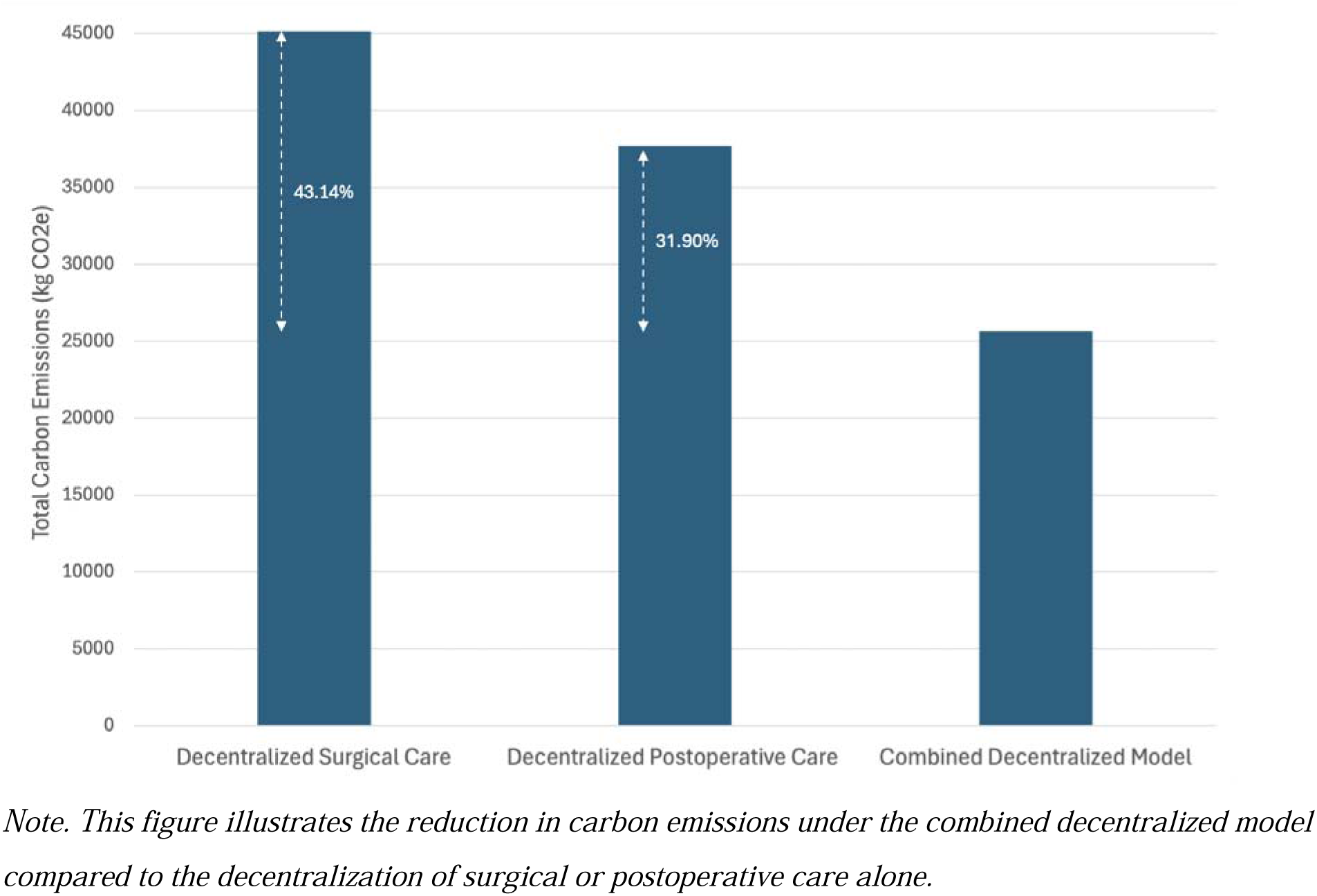
Comparison of the Combined vs Individual Decentralization Models

**Figure 7.**
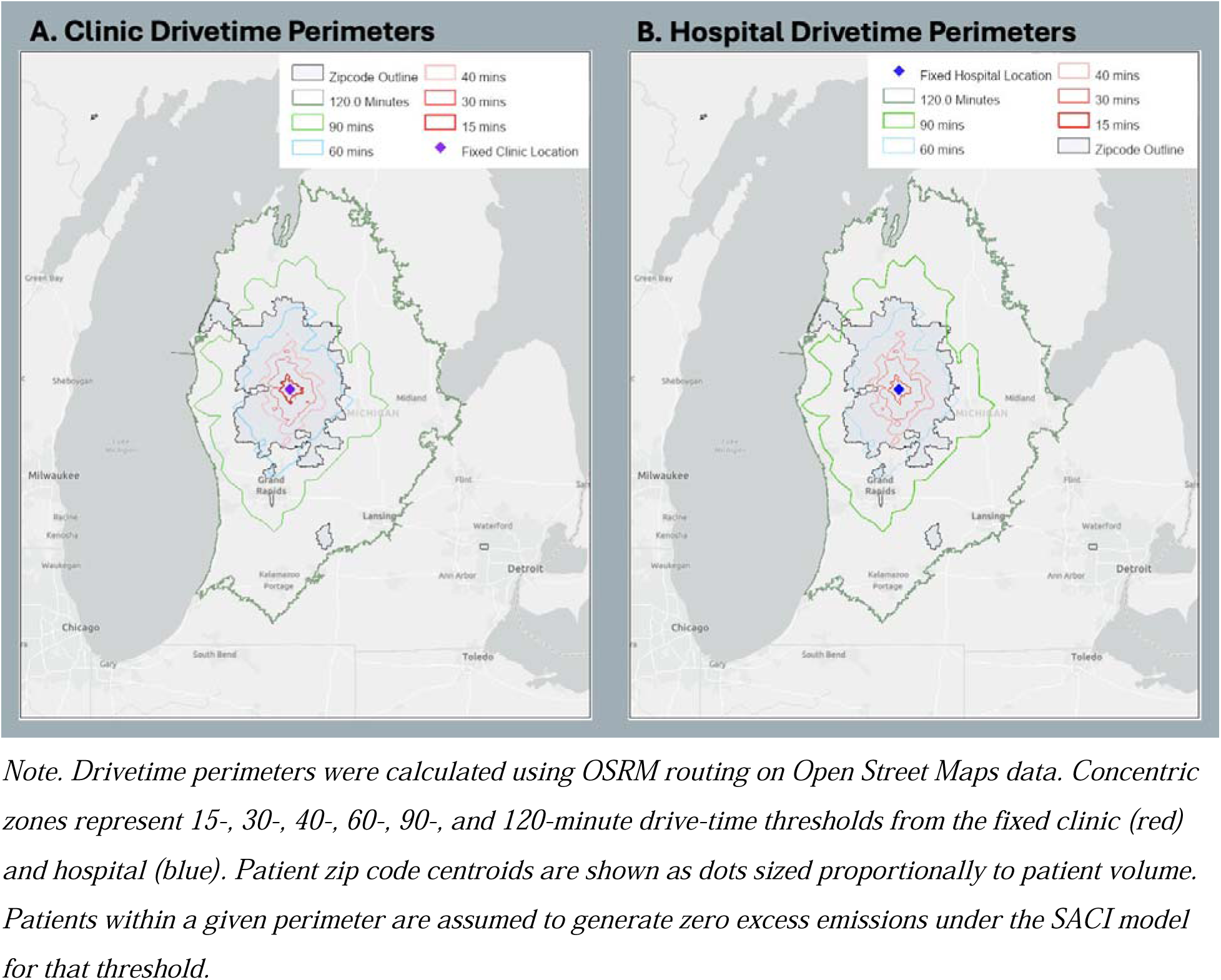
SACI Analysis: Drivetime Perimeters from Fixed Clinic and Hospital Locations.

**Figure 8.**
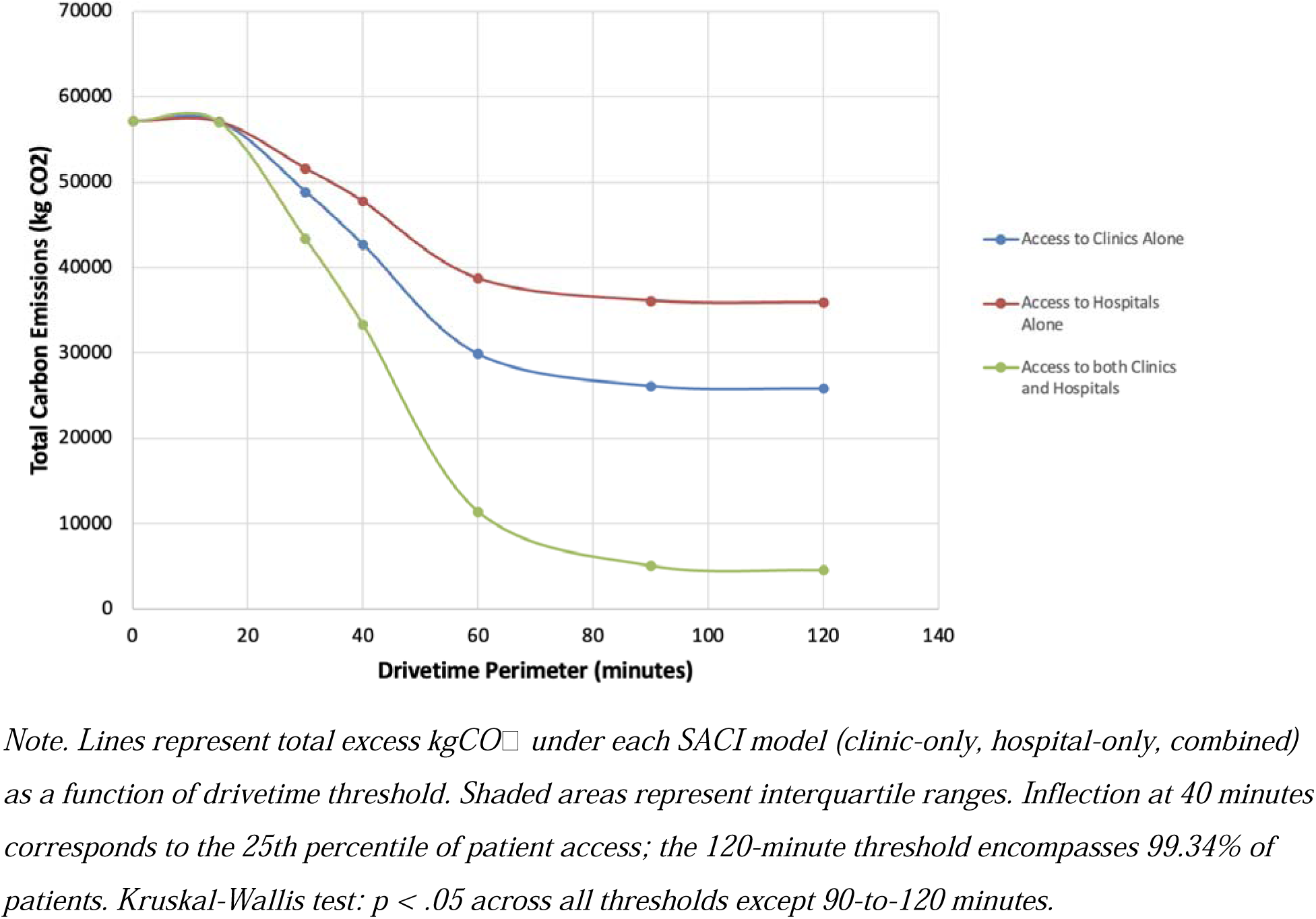
SACI: Impact of Timely Access on Total Carbon Emissions.

The CESR heatmap demonstrated a region of configurations with minimal gradient values beyond the inflection points of 7 hospitals and 9 clinics, indicating diminishing returns from further facility addition (Figure A2).

### SACI

At timeliness thresholds of 15, 30, 40, 60, 90, and 120 minutes, 164 (17.9%), 466 (50.87%), 604 (65.94%), 856 (93.45%), 908 (99.13%), and 910 (99.34%) patients, respectively, had access to surgical care within the specified drive time. Assuming access was achieved at each threshold, excess carbon emissions decreased from 57,054.62 kgCO (median: 59.20; IQR: 32.31-81.87 kgCO) at 15 minutes to 4,584.93 kgCO (median: 0; IQR: 0-0 kgCO) at 120 minutes (Table 4). An inflection point was observed at the 40-minute threshold (corresponding to the 25th percentile of access), at which 65.94% of patients lived within the perimeter, resulting in excess emissions of 33,324.35 kgCO . Timeliness thresholds beyond 60 minutes extended beyond the contiguous zip code region. The reduction in excess emissions across thresholds was statistically significant (p<0.05) for all increments except the step from 90 to 120 minutes. Achieving two-hour access to clinic services (120-minute threshold, clinics only) reduced total excess emissions by 54.7%, from 57,167.62 kgCO to 25,847.63 kgCO .

**Table 4.** SACI: Impact of Timely Access on Carbon Emissions.

| Drivetime<br>Perimeter<br>(mins) | Timely Access to Clinics Alone |  |  | Timely Access to Hospitals Alone |  |  | Timely Access to Both Clinics and<br>Hospitals |  |  |
| --- | --- | --- | --- | --- | --- | --- | --- | --- | --- |
|  | Total<br>Carbon<br>Emissions<br>*<br>(kgCO<sub>2</sub>) | Median<br>(IQR) | N (%)<br>within<br>Perimeter | Total<br>Carbon<br>Emissions*<br>(kgCO<sub>2</sub>) | Median<br>(IQR) | N (%)<br>within<br>Perimeter | Total<br>Carbon<br>Emissions*<br>(kgCO<sub>2</sub>) | Median<br>(IQR) | N (%)<br>within<br>Perimeter |
| **Centralized**<br>**Model** | 57,167.62 | 59.20<br>(32.31–<br>81.87) | N/A | 57,167.62 | 59.20<br>(32.31–<br>81.87) | N/A | 57,167.62 | 59.20<br>(32.31–<br>81.87) | N/A |
| **15** | 57,118.60 | 59.20<br>(32.31–<br>81.87) | 164<br>(17.90%) | 57,103.63 | 59.20<br>(32.31–<br>81.87) | 164<br>(17.90%) | 57,054.62 | 59.20<br>(32.31–<br>81.87) | 164<br>(17.90%) |
| **30** | 48,922.79 | 20.63<br>(16.11–<br>79.54) | 466<br>(50.87%) | 51,633.13 | 40.97<br>(18.81–<br>79.54) | 466<br>(50.87%) | 43,388.29 | 0 (0–79.54) | 466<br>(50.87%) |
| **40** | 42,731.06 | 20.63<br>(16.12–<br>65.48) | 620<br>(67.69%) | 47,760.90 | 34.38<br>(17.19–<br>68.68) | 604<br>(65.94%) | 33,324.35 | 0 (0–65.48) | 604<br>(65.94%) |

| Drivetime Perimeter (mins) | Timely Access to Clinics Alone |  |  | Timely Access to Hospitals Alone |  |  | Timely Access to Both Clinics and Hospitals |  |  |
| --- | --- | --- | --- | --- | --- | --- | --- | --- | --- |
|  | Total Carbon Emissions* (kgCO<sub>2</sub>) | Median (IQR) | N (%) within Perimeter | Total Carbon Emissions* (kgCO<sub>2</sub>) | Median (IQR) | N (%) within Perimeter | Total Carbon Emissions* (kgCO<sub>2</sub>) | Median (IQR) | N (%) within Perimeter |
| 60 | 29,899.54 | 20.63 (16.11–32.81) | 856 (93.45%) | 38,716.06 | 32.39 (16.94–42.87) | 856 (93.45%) | 11,447.98 | 0 (0–0) | 856 (93.45%) |
| 90 | 26,155.58 | 20.63 (16.11–32.81) | 908 (99.13%) | 36,110.06 | 32.39 (16.94–41.02) | 908 (99.13%) | 5,098.03 | 0 (0–0) | 908 (99.13%) |
| 120 | 25,847.63 | 20.63 (16.12–32.81) | 910 (99.34%) | 35,904.91 | 32.39 (16.94–41.02) | 910 (99.34%) | 4,584.93 | 0 (0–0) | 910 (99.34%) |

*Note. See Table A2 for sensitivity analysis accounting for patients lost to follow-up. \*Total carbon emissions include emissions from the initial surgical procedure and all postoperative visits. Total carbon emissions (CE) include emissions from the initial surgical procedure and all postoperative visits. Excess emissions represent kgCO<sub>2</sub> attributable to patients residing outside the relevant drivetime perimeter, under the assumption that patients within the perimeter require zero travel. A statistically significant reduction across timeliness thresholds was detected by Kruskal-Wallis test ( $p < .05$ ) for all increments except the step from 90 to 120 minutes. N (%) within = number and percentage of patients residing within the specified drivetime perimeter from the relevant facility type.*

### Sensitivity Analysis

A sensitivity analysis was conducted to evaluate the impact of variations in key parameters, such as the number of follow-up visits and assumptions about vehicle fuel efficiency. Adjusting the total number of follow-up visits from 1352 to 916 (1 post-op visit per patient) resulted in a 25.83% reduction (or 14768.39 kgCO), while improving vehicle fuel efficiency decreased emissions by 54.03% (or 30888.96 kgCO) and 57.12% (or 32656.23 kgCO), corresponding to the most efficient hybrid (Toyota Prius) and electric (Tesla Model 3) vehicles driven in Michigan, respectively. These results indicate that the model’s conclusions are robust across a range of plausible scenarios.

## Discussion

This study evaluates the carbon impact of decentralizing surgical care, specifically for cataract surgeries. The results demonstrate that centralized care models impose environmental costs due to increased travel, with the total emissions for all procedures and follow-ups under the current model reaching 57,167.62 kgCO . In contrast, decentralizing follow-up care through hypothetical clinic locations led to emissions reductions of up to 34.07%, or 19,475.02 kgCO, at optimal decentralization points. These findings suggest that decentralizing care could yield significant environmental benefits while improving timely access to surgical services.

The predominant approach to reducing healthcare’s carbon footprint has focused on hospital energy efficiency, pharmaceutical supply chains, and high-impact clinical inputs such as volatile anesthetic agents [13,14]. Comparatively little attention has been paid to care delivery architecture—the geographic distribution and organizational structure of services—as an environmental determinant. The OECD has explicitly identified transforming healthcare delivery as a key decarbonization pathway, yet empirical frameworks for doing so at the service-planning level remain scarce [15]. By demonstrating that the spatial configuration of surgical facilities is a quantifiable driver of travel-related emissions, this study provides an actionable, evidence-based lens through which planners can evaluate the environmental consequences of care organization decisions.

Centralized healthcare delivery models have been widely adopted in their perceived efficiency in consolidating expertise, resources, and advanced technologies. These models aim to improve patient outcomes through specialized care and streamlined service delivery. However, the broader implications of centralization to access to care, particularly for rural populations, are often overlooked. Centralized care disproportionately impacts patients who live far from these centers, imposing significant travel burdens, delays in access, and financial costs [16]. Although the facility and area described in this study was not formally designated a ‘centre of excellence’, it was the sole provider of ophthalmologic surgery within the catchment area, effectively functioning as a centralized facility and producing similar burdens on patients. Cataract surgeries were selected for this study as a representative model due to their defined care cascade, discrete and predictable care pathway, and reliance on in-person postoperative evaluations. These characteristics make ophthalmologic procedures well-suited for exploring the interplay between care delivery models and environmental impacts. The two clinics for postoperative follow-up form part of the centralized service. Our findings demonstrate that decentralizing follow-up care through additional clinic locations could reduce emissions by up to 39.52%, while decentralizing surgical care at the hospital level could reduce emissions by up to 29.09%.

A central objection to surgical decentralization is that distributing procedures across more facilities risks diluting volume per site, with adverse effects on quality. This concern is well-founded for high-acuity complex procedures such as pancreatectomy, oesophagectomy, and complex cancer surgery, where robust volume-outcome evidence supports centralization [17]. For cataract surgery, the evidence is different: quality improvements plateau at relatively low surgeon volumes (approximately 200–350 cases per year), and facility-level volume does not independently predict adverse outcomes [18]. This makes cataract surgery, and by extension a wide range of high-volume, lower-acuity elective procedures, an appropriate and clinically defensible target for decentralized delivery. Any proposal to decentralize higher-acuity procedures would require specific analysis of the volume-quality tradeoff for that procedure.

This study adds to the prevailing narrative around making healthcare more ‘green’ which focuses largely on consumption of hospital facilities or the broader climate narrative around improving vehicle fuel efficiency as a means of decreasing emissions. Without accounting for the carbon impact of patient travel, centralization risks undermining broader sustainability goals, particularly in the context of global efforts to reduce healthcare-related emissions [11]. Rural populations simultaneously bear the heaviest burden of inadequate surgical access and generate the highest per-trip healthcare travel emissions [12,19]. Climate change disproportionately affects rural communities through extreme weather events, agricultural disruption, and infrastructure vulnerability, compounding pre-existing socioeconomic disadvantage [20]. Healthcare systems that impose long-distance travel on rural patients are simultaneously extracting access equity from the patients themselves while extracting environmental capacity from communities least equipped to absorb the cost. Geospatial adjustments in facility distribution, when paired with existing efforts to change transportation types, offer a synergistic strategy for reducing emissions, particularly for follow-up care [8,3]. However, we found that decentralization of care, particularly follow-up care, can have a more substantial effect on overall vehicle emission reduction. This suggests that systemic reorganization of healthcare delivery, including geospatial restructuring, could have broader environmental and patient-centred benefits.

Population health access indicators play a pivotal role in bridging healthcare and climate policy. Indicators like the LCoGS two-hour access benchmark are traditionally assessed through a population health lens, evaluating how distance and travel time affect patient outcomes and timely intervention. However, this study demonstrates that these access indicators also have environmental implications. When populations are unable to access care within the two-hour threshold, longer travel distances increase the carbon footprint of healthcare delivery. We proposed the SACI indicator as a practical way to integrate access to care and climate impact for surgical care. While this study is grounded in a single rural Michigan practice, the SACI framework and k-means facility optimization approach are procedure-agnostic and geographically transferable. The analytical pipeline could be applied to any elective surgical procedure with a defined care cascade including colonoscopy, hernia repair, or orthopaedic procedures, and in any setting with geospatial and electronic medical record data. This is particularly relevant for low-and middle-income countries developing national surgical plans (NSPs) as part of the global surgery agenda. As these plans expand the surgical facility platform to meet SDG 3.8 targets, spatial planning decisions will lock in decades of access and emissions outcomes. Incorporating SACI into NSP monitoring frameworks would allow planners to simultaneously optimize for clinical access and environmental impact through a dual mandate not currently reflected in any existing global surgery indicator [21].

### Limitations

This study is not without limitations. First, the absence of granular vehicle data required assumptions about the most commonly used vehicles in the state. However, we include sensitivity analyses to provide lower-bound estimates on the carbon savings from optimal healthcare center placement, assuming everybody drove the most fuel-efficient vehicles available in Michigan at the time of this study. Secondly, our use of population-weighted centroids to approximate travel distances may not fully reflect individual travel patterns. Although the k-means clustering approach approximates optimal facility placement, it does not guarantee a global minimum for emissions. Finally the study focuses on the climate benefits of decentralization without considering the broader impact on specialized infrastructure and expertise only available in centralized facilities; these domains are adequately addressed in other studies of centralization. Additionally, we did not conduct a life cycle assessment (LCA) of the environmental impact of building additional facilities, which could partially offset the emissions savings. Future research should balance environmental considerations with known benefits of centralization, including clinical safety, efficiency, and resource optimization. The findings here, however, lay the groundwork for a comprehensive CBA approach by contributing a climate criterion to decision-making frameworks.

### Policy implications

1. **Optimize the number of facilities**: Our analysis identified a local minimum configuration of additional clinics and hospitals to achieve significant emission reductions and improved access to care, with potential flexibility depending on population density and demand. This model should be considered when planning for the expansion of the service delivery platform. While this model did not account for the potential of increasing facilities to induce demand and increase patient visits), future implementation of the model would require evaluation and patient behavior changes.
2. **Incorporate climate metrics into access monitoring**: Population health access indicators, like the LCoGS two-hour threshold, should explicitly account for the environmental impact of unmet clinical access standards. Failing to meet these thresholds results in adverse health outcomes and environmental harm.
3. 3. **Balance decentralization and centralization by procedure type**: Policymakers should balance the known clinical benefits of centralized care with the environmental and access-related benefits of decentralization, tailoring solutions to local contexts. Health policies aiming at evaluating access to and quality of care, can use our indicator, and any access indicator with an inherent travel time, to evaluate the climate impact and gains of access to care.
4. 4. **Integrate spatial equity into climate-health policy**: Decentralizing surgical care in rural settings simultaneously addresses the access equity deficit and the disproportionate travel carbon burden borne by rural communities. Climate-health policy frameworks should recognize this dual benefit

## Conclusion

Decentralizing surgical care delivery presents a significant and underrecognized opportunity to reduce the carbon footprint of healthcare, particularly in rural areas. This study demonstrates that the spatial organization of surgical services is a quantifiable, modifiable driver of travel-related emissions, reducible by up to 40% through evidence-based facility planning. The SACI metric provides a novel framework for integrating environmental sustainability into surgical care planning, and the analytical pipeline is transferable to other procedures, settings, and national contexts. Meeting global surgical access benchmarks and achieving healthcare sustainability goals are not competing imperatives.

## Data Availability

All data produced in the present study are available upon reasonable request to the authors

## Funding

None

## Conflicts of interest

The authors declare no conflicts of interest.

## Ethics

Not Human Subjects Research- Harvard Longwood Campus IRB, Protocol #IRB24-1130, September 6, 2024.

## Author Contributions

Conceptualization: GYH, RC, MS

Methodology: GYH, RR, TW, MS

Formal analysis: GYH, RR, TW

Data curation: RR, GYH

Writing - original draft: GYH, RR

Writing - review & editing: GYH, RR, TW, RC, MS

Supervision: MS

Final approval: all authors

## Notes

### Competing Interest Statement

The authors have declared no competing interest.

### Author Declarations

Not Human Subjects Research- Harvard Longwood Campus IRB, Protocol #IRB24-1130, September 6, 2024.

