## Appendix 1 for "Barriers to surgical care delivery are harming our planet: a case for decentralized provider services"

#### Appendix 1A. Distribution of Distinct Zipcodes, Patient Counts, and Total Post-op Visits.

| **Zipcode** | **# of Patients** | **Total # of Postops** | **Distance to Clinic (in km)** |
| --- | --- | --- | --- |
| 49307 | 164 | 240 | 0.2745 |
| 49338 | 18 | 28 | 13.7094 |
| 49342 | 19 | 31 | 18.2284 |
| 49320 | 6 | 8 | 21.41 |
| 49346 | 72 | 111 | 21.7676 |
| 49639 | 34 | 50 | 23.1077 |
| 49336 | 36 | 53 | 25.2889 |
| 49677 | 100 | 145 | 27.5283 |
| 49332 | 51 | 78 | 27.5326 |
| 49305 | 16 | 21 | 33.3614 |
| 49623 | 12 | 15 | 38.5883 |
| 49340 | 6 | 7 | 39.6892 |
| 49309 | 15 | 26 | 39.7338 |
| 49329 | 59 | 89 | 40.3395 |
| 49631 | 85 | 121 | 43.9115 |
| 49349 | 27 | 38 | 44.1526 |
| 49339 | 13 | 19 | 46.1583 |
| 49679 | 13 | 20 | 47.631 |
| 49655 | 17 | 25 | 48.592 |
| 49642 | 7 | 9 | 49.0494 |
| 49343 | 3 | 4 | 51.3457 |
| 49337 | 26 | 37 | 53.5025 |
| 49304 | 29 | 46 | 55.0721 |
| 48850 | 12 | 18 | 55.1354 |
| 49310 | 1 | 1 | 56.0838 |
| 49688 | 9 | 12 | 56.3289 |
| 49656 | 12 | 17 | 57.6243 |
| 48893 | 1 | 2 | 59.1449 |
| 48886 | 1 | 1 | 62.0436 |
| 49347 | 7 | 11 | 64.1472 |
| 48632 | 4 | 7 | 65.9924 |
| 49412 | 2 | 3 | 66.0699 |
| 49327 | 2 | 3 | 68.499 |
| 49330 | 1 | 2 | 74.4119 |
| 49601 | 4 | 5 | 74.4326 |
| 49665 | 7 | 10 | 78.2592 |
| 48888 | 3 | 5 | 80.7797 |
| 49321 | 2 | 6 | 83.7611 |
| 49618 | 1 | 2 | 83.9619 |
| 49644 | 6 | 8 | 84.415 |
| 48838 | 3 | 4 | 89.7035 |
| 49548 | 2 | 2 | 101.0573 |
| 49660 | 2 | 3 | 137.9713 |
| 49073 | 1 | 1 | 176.8684 |
| 48377 | 1 | 2 | 296.566 |
| 49868 | 2 | 3 | 416.5041 |
| 49886 | 2 | 3 | 567.4402 |

#### Appendix XXA. Sensitivity Analysis of Total Carbon Emissions: Accounting for Patients Lost to Follow-Up in Centralized vs. Decentralized Post-Operative Care

*Please note that the total carbon emissions include emissions from the initial operative procedure and post-operative visits.

**Sensitivity analysis was performed to account for the 6 patients lost to follow-up, using the mean number of post-operative visits (1.48) instead of 0. One-tailed Wilcoxon Signed-Rank Test was performed, given the skewed dataset. A p-value of 0.00195 suggests that the sensitivity analysis leads to a statistically significant increase in carbon emissions.

| **Number of Clinic Locations (n + 1)** | **Total Carbon Emissions*** | **Median (IQR) of Total Carbon Emissions** | **Sensitivity Analysis**** | **Difference** |
| --- | --- | --- | --- | --- |
| Fixed Hospital (1) | 57167.62 | 59.20 (49.55) | 57325.81 | 158.19 |
| 2 | 50392.16 | 48.51 (33.31) | 50530.04 | 137.88 |
| 3 | 46179.77 | 40.89 (29.16) | 46269.19 | 89.42 |
| 5 | 42502.71 | 40.89 (25.94) | 42594.89 | 92.18 |
| 7 | 39317.55 | 37.45 (25.92) | 39386.24 | 68.69 |
| 9 | 37692.60 | 37.47 (23.93) | 37775.12 | 82.52 |
| 11 | 36896.15 | 36.21 (30.64) | 36962.47 | 66.32 |
| 13 | 36365.68 | 38.61 (21.06) | 36448.20 | 82.52 |
| 15 | 34642.93 | 33.63 (21.47) | 34678.52 | 35.59 |
| 17 | 34577.03 | 34.75 (21.36) | 34634.65 | 57.62 |

**Appendix Figure XXA. Impact of Decentralized Surgical and Postoperative Care on Total Carbon Emissions.** This heatmap illustrates total carbon emissions across various configurations of hospital and clinic locations. Gray regions between shaded squares indicate permutations without primary data. The blue dashed outline (9 clinics; 7 hospitals) shows the combination of inflection points.

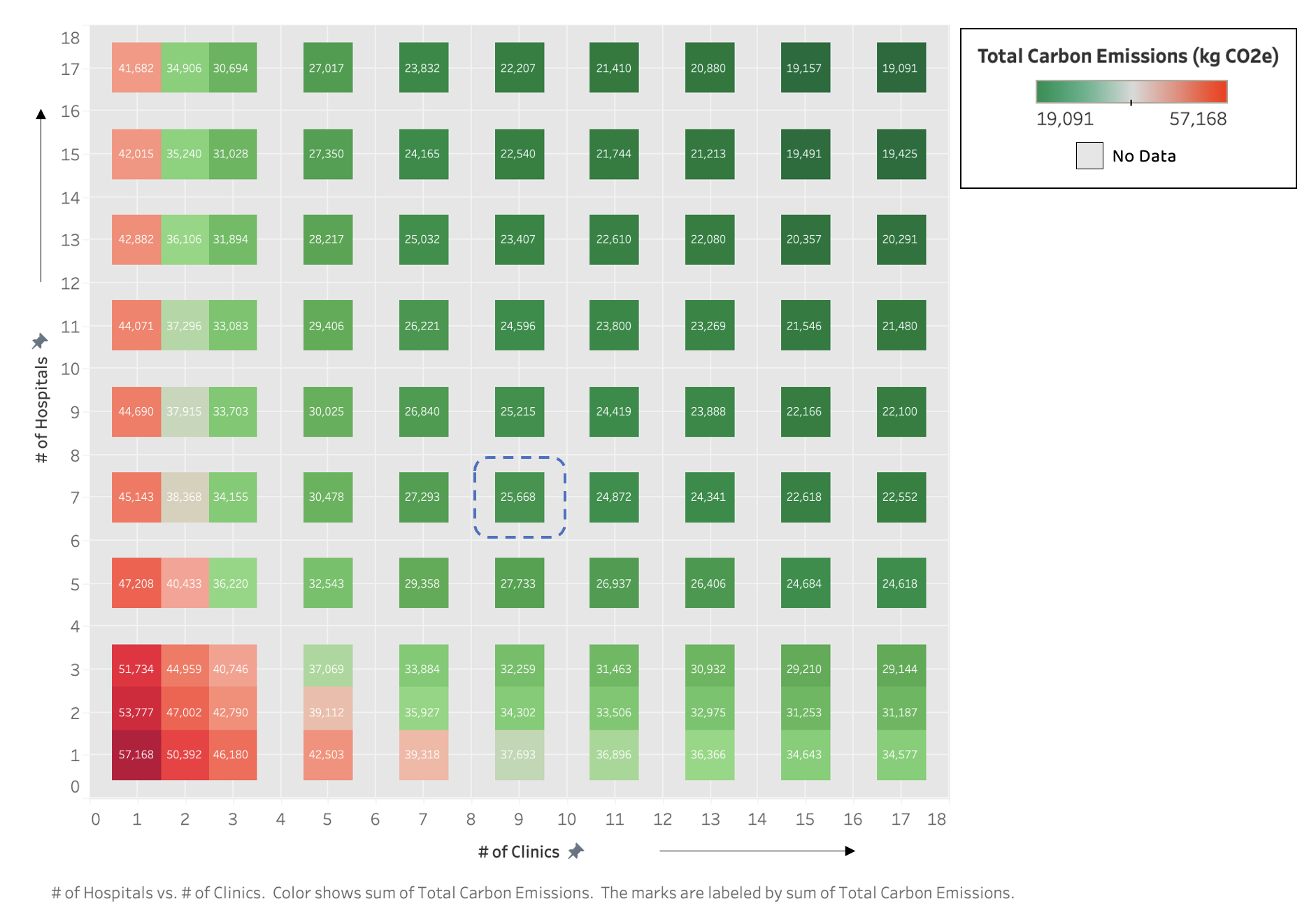

**Appendix XXA. Gradient Analysis of Total Carbon Emission Reduction.**

This heatmap shows the rate of change in total carbon emission reduction, calculated as the gradient magnitude: the square root of the sum of squared first derivatives along the x- and y-axes. It evaluates variations across different configurations of hospital and clinic locations. Gray regions indicate permutations without primary data, while the blue outline highlights configurations with minimal gradient values.

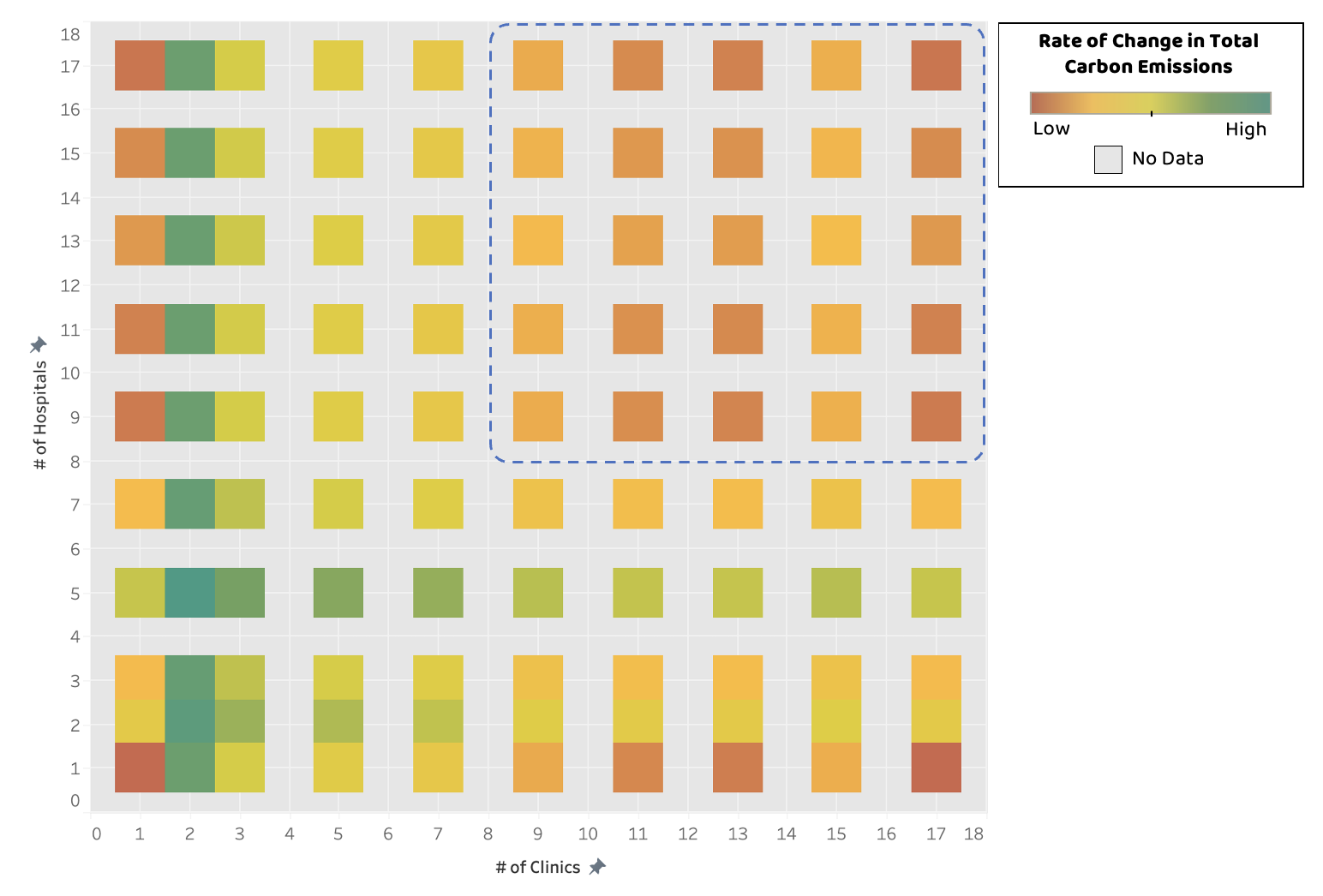

#### Figure XXA. Additional Hospital Locations. The following maps show the location of the fixed hospital and n - 1 additional hospital locations, for a total of n hospital locations.

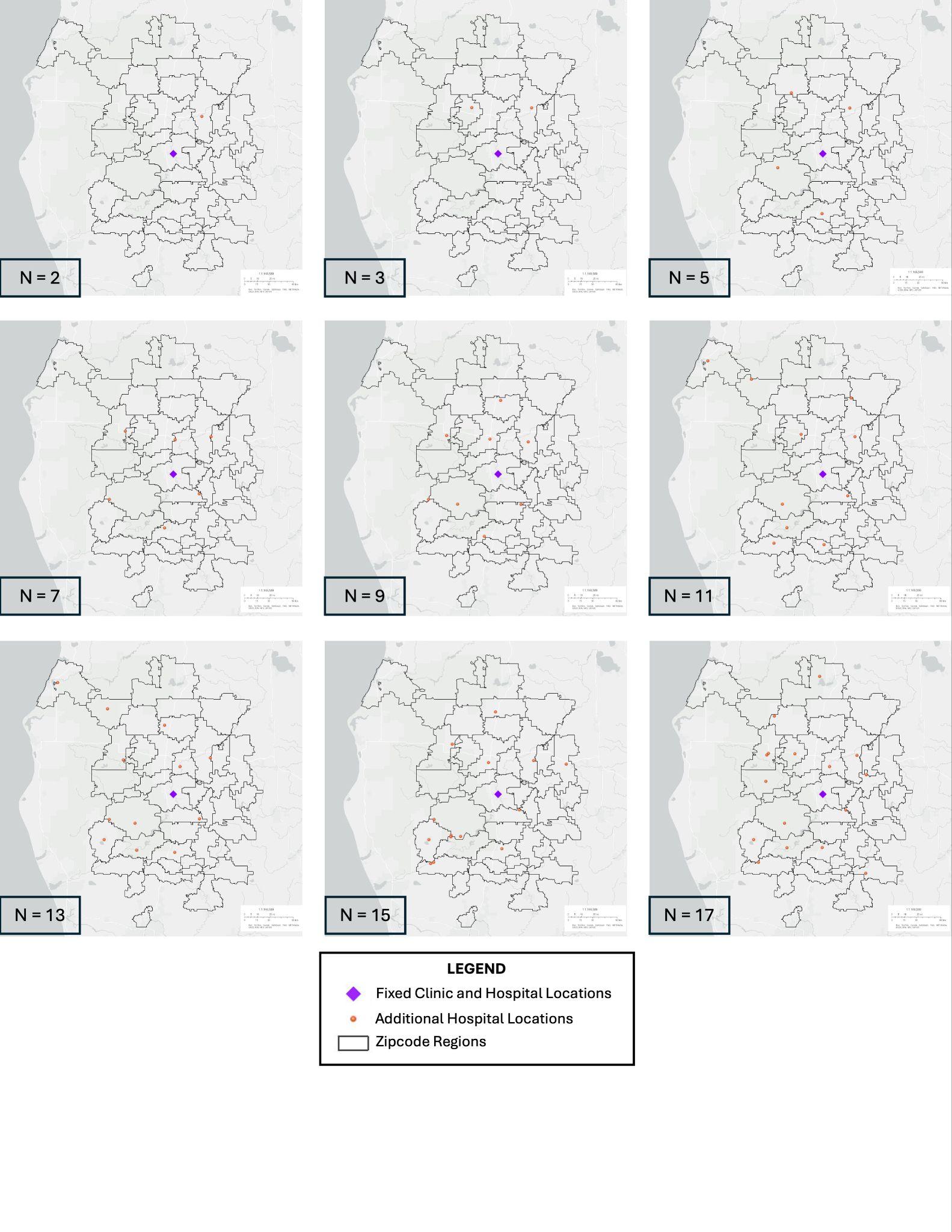

#### Figure XXA. Additional Clinic Locations. The following maps show the location of the fixed clinic and n - 1 additional clinic locations, for a total of n clinic locations.

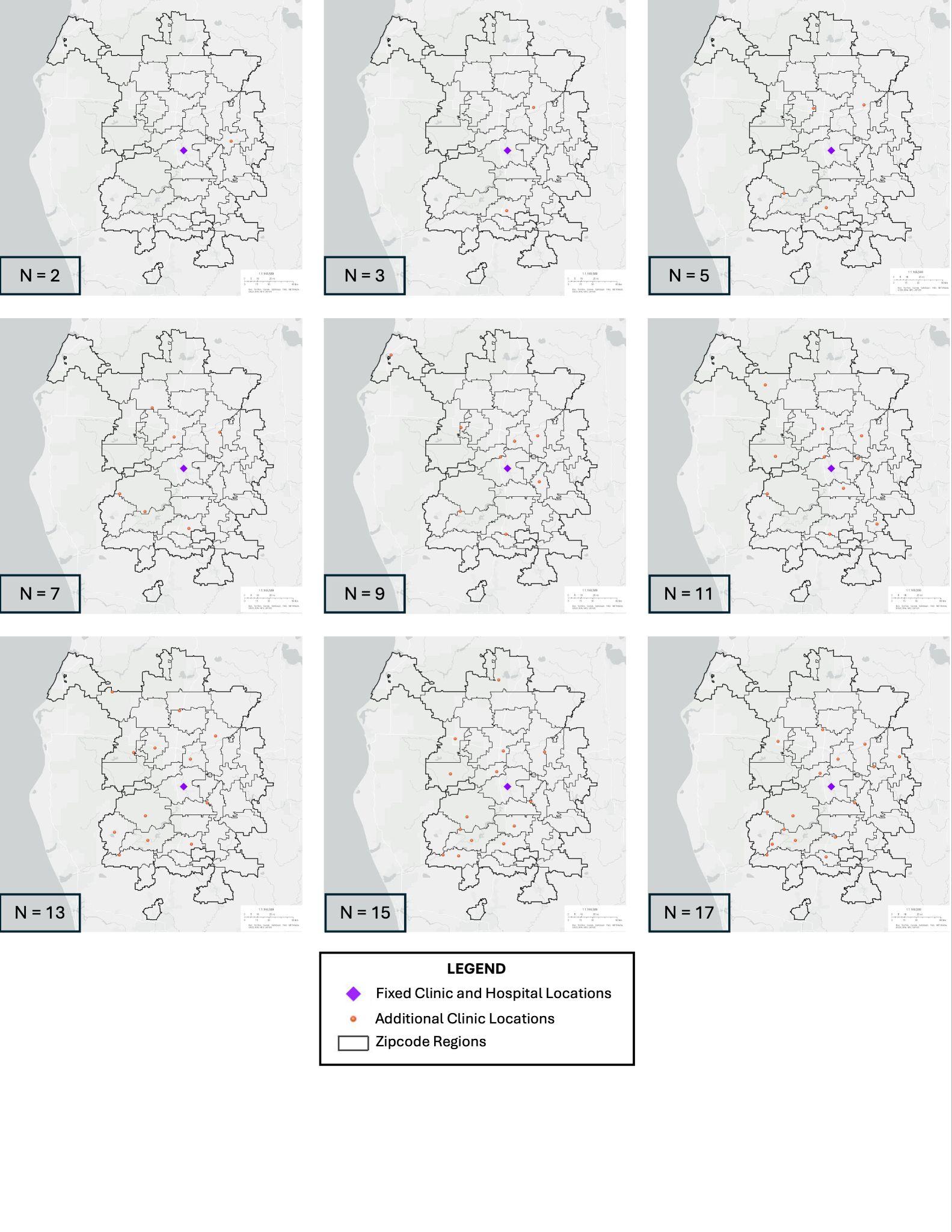
